# Impact analysis of infant antibiotic exposure on the burden of asthma: a simulation modeling study

**DOI:** 10.1101/2024.07.19.24310721

**Authors:** Tae Yoon Lee, John Petkau, Ariana Saatchi, Fawziah Marra, Stuart E Turvey, Hannah Lishman, David M Patrick, Jacquelyn J Cragg, Kate M Johnson, Mohsen Sadatsafavi

## Abstract

**Background:** Infant antibiotic use is associated with increased risk of asthma. We examined the population impact of antibiotic exposure in the first year of life on the burden of pediatric asthma in British Columbia, Canada, using simulation modeling.

**Methods:** We performed a Bayesian meta-analysis of empirical studies to construct dose-response equations between antibiotic exposure in the first year of life and pediatric (<19 years of age) asthma. We used administrative health data to document trends in infant (< 1 year of age) antibiotic use in British Columbia during 2001 and 2018 (the study period). An independently developed microsimulation model of asthma was utilized to estimate asthma-related outcomes under three scenarios pertaining to the trends in antibiotic use during the study period: 1) observed trends, 2) flat trend in which the prescription rate remained at the 2001 value, and 3) intermediate trends midway between these two. We reported cumulative person-years with asthma, cumulative asthma incidence, and cumulative asthma exacerbations among the pediatric population during the study period.

**Results:** There were 773,160 live births during the study period, with an average antibiotic prescription rate of 523 per 1,000 infants in the first year of life. The prescription rate decreased by 71.5% during the study period. In Scenario 1, there were 1,982,861 person-years with asthma, 183,392 asthma incident cases, and 383,072 exacerbations. Had the antibiotic exposure remained at the 2001 values (Scenario 2), there would have been additional 37,213 person-years with asthma, 10,053 asthma incident cases, and 23,280 exacerbations. Had the decline been half of the observed trend (Scenario 3), there would have been additional 20,318 person-years with asthma, 5,486 asthma incident cases, and 12,728 exacerbations. At least 80% of the excess burden in each outcome was attributable to the younger pediatric population of <10 years of age.

**Conclusions:** The decline in infant antibiotic exposure has resulted in a substantial reduction in the burden of asthma in British Columbia. Such benefits should be considered when evaluating the value proposition of initiatives aimed at reducing unnecessary antibiotic exposure in early life.

## 1 Introduction

Overuse of antibiotics is a global health problem, with approximately 30% of outpatient antibiotic prescriptions estimated to be unnecessary (1). Many health jurisdictions across the world have undertaken initiatives, such as launching antimicrobial stewardship programs, to reduce unnecessary use of antibiotics. A recent systematic review of 52 studies reporting on such stewardship programs found an overall 10% attributable reduction in antibiotic prescriptions (2). However, it is not clear to what extent such a reduction will impact the health of the population. Understanding the population and policy implications of interventions is necessary for evidence-informed and efficient decision-making (3).

A prime example of the benefit of reduced antibiotic exposure is the potential reduction in the burden of asthma. Asthma is a common chronic inflammatory airway condition characterized by airflow restriction, heightened airway responsiveness, and structural alterations in the air passages (4). Asthma continues to present a public health challenge, affecting over 262 million people globally (5,6). Numerous studies have shown asthma to result in premature mortality, diminished quality of life, and substantial economic burden (7).

Growing evidence shows a link between infant antibiotic use and childhood asthma development (8). This link can be attributed to the deleterious impact of antibiotics on the maturation of the infant gut microbiome, the composition of which plays a crucial role in healthy immunological development. Disruption of this process can lead to hyperinflammatory immune responses later in childhood and the development of allergic diseases such as asthma in later life (9–11). Therefore, it is plausible that the concomitant reduction in antibiotic use and the incidence of asthma observed in many jurisdictions may be related (11–14). For example, a 73% reduction (from 868 to 236 prescriptions per 1,000 population) in antibiotic use among infants was paralleled with a 41% reduction in asthma incidence (from 29 to 17 incident cases per 1,000 population) among children aged 1–4 years between 2000 and 2018 in British Columbia (15). A concern about such associations is potential confounding by indication or reverse causation due to respiratory infection since antibiotics are indicated for respiratory infections, and respiratory infections are a risk factor for asthma (16). However, the association with asthma remains significant after adjusting for respiratory infections, and is also observed when restricting the outcome to non-respiratory indications (11). Moreover, at the ecological level, dramatic falls in childhood asthma rates following reductions in antibiotic use were not preceded by concomitant changes in the population prevalence of respiratory infections (11). The established pathway through missing taxa, altered metabolites and T cell development adds biologic plausibility for a causal association (17). Putting these together, preventing the early steps in asthma pathogenesis during infancy through reduction in antibiotic exposure presents a unique opportunity for asthma prevention (18).

Given the strength of evidence towards the causal association between antibiotic exposure and risk of childhood asthma, we sought to investigate the policy implications of reducing unnecessary antibiotic use. Focusing on British Columbia (BC), a Canadian province with a 2022 population of 5.4 million (19), our study aimed to address the question: “*What would the burden of asthma have been, had infant antibiotic exposure been different than observed?”* Specifically, the objective of our study was to project population-level asthma-related outcomes under different counter-factual scenarios related to recent declining trends in antibiotic exposure among infants in BC and then to estimate the concomitant changes in the burden of asthma that can be attributed to such trends.

## 2 Materials and methods

This study comprised of three major steps to achieve the objective: 1) ecological trend analysis of infant antibiotic exposure in BC, 2) meta-analysis of the dose-response relationship between infant antibiotic use and risk of childhood asthma, and 3) simulation modeling to quantify asthma burden under counterfactual scenarios regarding infant antibiotic exposure. The study period for which antibiotic exposure was quantified and asthma outcomes were modeled was 2001–2018.

All the analyses were conducted using R (version: 4.3.0) (20), Julia (version: 1.9.0) (21), and Stan (2.21) (22). The aggregated data and code used for the analyses are available in the GitHub repository: https://github.com/tyhlee/ImpactInfantAbxAsthma. This study was approved by the institutional review board of the University of British Columbia, Vancouver (H09–00650).

### 2.1 Ecological trend analysis of antibiotic exposure among infants in BC

We analyzed antibiotic use among infants (< 1 year of age) using the population-based administrative health databases of BC from January 2001 (earliest year allowed in the simulation model) to December 2018 (last year of data availability for this study). In particular, the PharmaNet database contains dispensed medication records across community pharmacies and hospital outpatient pharmacies for all residents of the province (23). Medications dispensed to inpatients within hospitals or emergency departments are not included within this database. However, such prescriptions are less likely to be misused (and thus are unlikely to be affected by any policy targeting antibiotic exposure reduction), and are typically short-term. Data were extracted, anonymized, and made available to researchers by the BCCDC (Do Bugs Need Drugs? (H09-00650)). All inferences, opinions and conclusions drawn in this study are those of the authors and do not reflect the opinions or policies of the Data Steward(s).

We computed the rate of infant antibiotic prescriptions per 1,000 infants using the population estimates from Statistics British Columbia (19). To estimate sex-specific trends, we fitted a negative binomial regression model for the log of the prescription rate, with calendar time and biological sex as covariates. An earlier study indicated a distinct decline in antibiotic use in 2005, coinciding with the introduction of a provincial antibiotic stewardship program (25). To account for this non-linearity, we also included main-effect and interaction-effect terms between time and an indicator for whether the year was after 2005. We measured the goodness-of-fit of the model with the percentage deviance explained (the higher deviance explained the better, analogous to R^2^ in an ordinary linear model, with values close to 100% justifying the use of regression-smoothed values instead of raw frequencies) (26).

### 2.2 Meta-analysis of the dose-response relationship between infant antibiotic use and childhood asthma prevalence

The analysis aimed to generate an equation that would map antibiotic exposure to asthma prevalence based on published information. We started with 63 studies that were identified in the systematic review by Duong et al. (8) which reported on the relationship between antibiotic use in early life and childhood asthma during the publication period between 1998 and 2021. We performed a further literature review (January 2021 to September 2023) to identify any relevant studies published afterwards (details of the search are provided in **Supplementary Materials Section 1**; 13 studies were identified).

Using the 76 identified studies, we estimated the dose-response relationship between infant antibiotic use and childhood asthma prevalence (in terms of odds ratio). We applied the following inclusion criteria to the identified studies: (i) the dose-response relationship was investigated as part of an original research (not synthesized from other studies), (ii) the timing of antibiotic use was in the first year of life, (iii) the age range for asthma diagnosis was narrower than 6 years (if asthma diagnosis is made in children of ages between 3 and 6, then the age range is 4 years) and (iv) the risk of bias score was low (the overall risk of bias less than 2 based on the risk of bias domains of ROBINS-I (27) following the guidelines by the Agency for Healthcare Research and Quality (28)).

We made several assumptions and simplifications to enable the quantitative pooling of results. Specifically, some studies reported the association for a single year of age, while others documented the association over an age bracket. Based on dedicated simulation studies (see **Supplementary Materials Section 2**), we concluded that the effect size reported across an age range of less than 6 years would provide a good approximation for the single-year effect at the mid-point of the age range (studies reporting on longer ranges were thus excluded, see criterion iii above). Second, we treated hazard ratios and odds ratios interchangeably, which is an acceptable approximation when event prevalence is low (29). Further, we assumed that the dose-response association was proportional on the logit scale (this assumption was qualitatively evaluated based on comparing the predicted versus observed association for each study). Next, we classified the number of antibiotic prescriptions (of any days supply) as 0, 1, 2, 3, 4, and 5+. As some studies reported the associations for the number of antibiotic prescriptions as a range, we used the following categorization by taking the lower bound of the range: 1–2 prescriptions were recategorized as 1, and 3–4 prescriptions were recategorized as 3. Lastly, the included studies did not report on the interaction effect with sex, and existing evidence on this topic is weak (30); correspondingly, we assumed that the effect of infant antibiotic use was not different between males and females.

We fitted a random-effects meta-regression model with age of asthma diagnosis and infant antibiotic use as covariates in a Bayesian framework (31) (details on the model, implementation in Stan, and prior specification are provided in **Supplementary Materials Section 3**). The final equation was of the form:

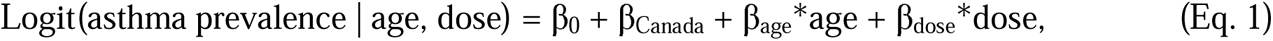

where age is the age of asthma diagnosis in years, dose is the number of antibiotic prescripts (0, 1, …, 4, 5+), β_0_, β_age_, β_dose_ are the fixed effects estimated by the meta-regression and β_Canada_ is the random effect corresponding to the single Canadian study in that analysis. This model specification enables pooling of all available evidence while also putting more weight on the single Canadian study.

### 2.3 Asthma policy simulation model

We used the Lifetime Exposures and Asthma outcomes Projection (LEAP) model, an asthma policy simulation model for Canada, to simulate counterfactual scenarios. Following the best practice recommendations in policy modeling (32), the LEAP model was developed, independently of this study, in collaboration with a steering committee of economic modelers, allergists, and respirologists across Canada. Model design, including important risk factors and the pathway of asthma, was informed from multiple rounds of Delphi processes to achieve consensus.

Details of this process and the model are reported elsewhere (33,34). In brief, the LEAP model is an open-population discrete-time microsimulation platform that simulates a virtual Canadian population and follows it from birth or immigration to emigration, death, or the end of the pre-specified time horizon. After initializing the population in the starting year (2001 in our case), simulated individuals enter the model by birth or immigration in subsequent years. In each year, the actions and behaviors of the individual are simulated based on pre-defined mathematical equations, and the attributes and disease characteristics of the individual are updated accordingly. LEAP models a multitude of population outcomes including asthma incidence, prevalence, and exacerbations.

The LEAP model has been calibrated to the Canadian population, mimicking the demographic and disease characteristics of the Canadian population. Asthma incidence and prevalence were modeled and calibrated using the Chronic Disease Registry (35), and asthma exacerbations were calibrated using the Hospital Morbidity Database (36). Further, we incorporated the equations for antibiotic exposure among infants and the effect of infant antibiotic exposure on asthma prevalence (developed in the previous sections) into the LEAP model and then calibrated the LEAP model. Of note, LEAP does not model the disease course of asthma for children under 3 years of age due to difficulty in reliably diagnosing asthma in this age group. Further details of the LEAP model are provided elsewhere (34).

### 2.4 Evaluation of counterfactual scenarios

To investigate the impact of the observed decline in antibiotic use in BC between 2001–2018, we compared the following ‘what-if’ scenarios. Scenario 1 was the base (factual) scenario using the smoothed trends (from the negative binomial regression) fitted to the observed data. Scenario 2 was a counterfactual scenario that modeled flat trends where the antibiotic prescription rate remained the same as the smoothed values in 2001. Scenario 3 was another counterfactual scenario that modeled a decline in the antibiotic prescription rate that was half of the smoothed trends (i.e., for any given year, the antibiotic prescription rate was the average of those in Scenarios 1 and 2).

The model was run separately for each scenario for the entire pediatric population (<19 years of age) of BC (approximately 1 million people in 2018) for 100 times. For each scenario, the following outcomes were recorded: total person-years with asthma (accounting for both the number of individuals with asthma and the amount of time each person with asthma spends during the study period), cumulative asthma incidence, and cumulative asthma exacerbations. We reported the average values of these measures across the 100 runs along with Monte Carlo (MC) standard deviations (SD). The results were provided by the following age groups (in years) as well: 3–4, 5–9, 10–14, and 15–18. Additionally, for each counterfactual scenario (Scenarios 2 and 3), differences and relative changes in each of the outcomes compared with the base scenario (Scenario 1) were recorded.

## 3 Results

### 3.1 Trends in antibiotic use

Over the period from 2001 to 2018, there were 773,160 infants born in BC. During the same period, 404,675 prescriptions of antibiotics for infants were recorded, corresponding to 523 prescriptions per 1,000 infants. Male infants received more prescriptions (586 per 1,000) than female infants (457 per 1,000). From 2001 to 2018, the number of female and male infants grew by 10.2% and 8.1%, respectively, whereas the prescription rate decreased by 71.2% for females and 71.6% for males, indicating a substantial decline in antibiotic exposure, as depicted in **Figure 1**. The negative binomial regression model fitted the data well with a goodness-of-fit of 99.3% deviance explained.

**Figure 1.**
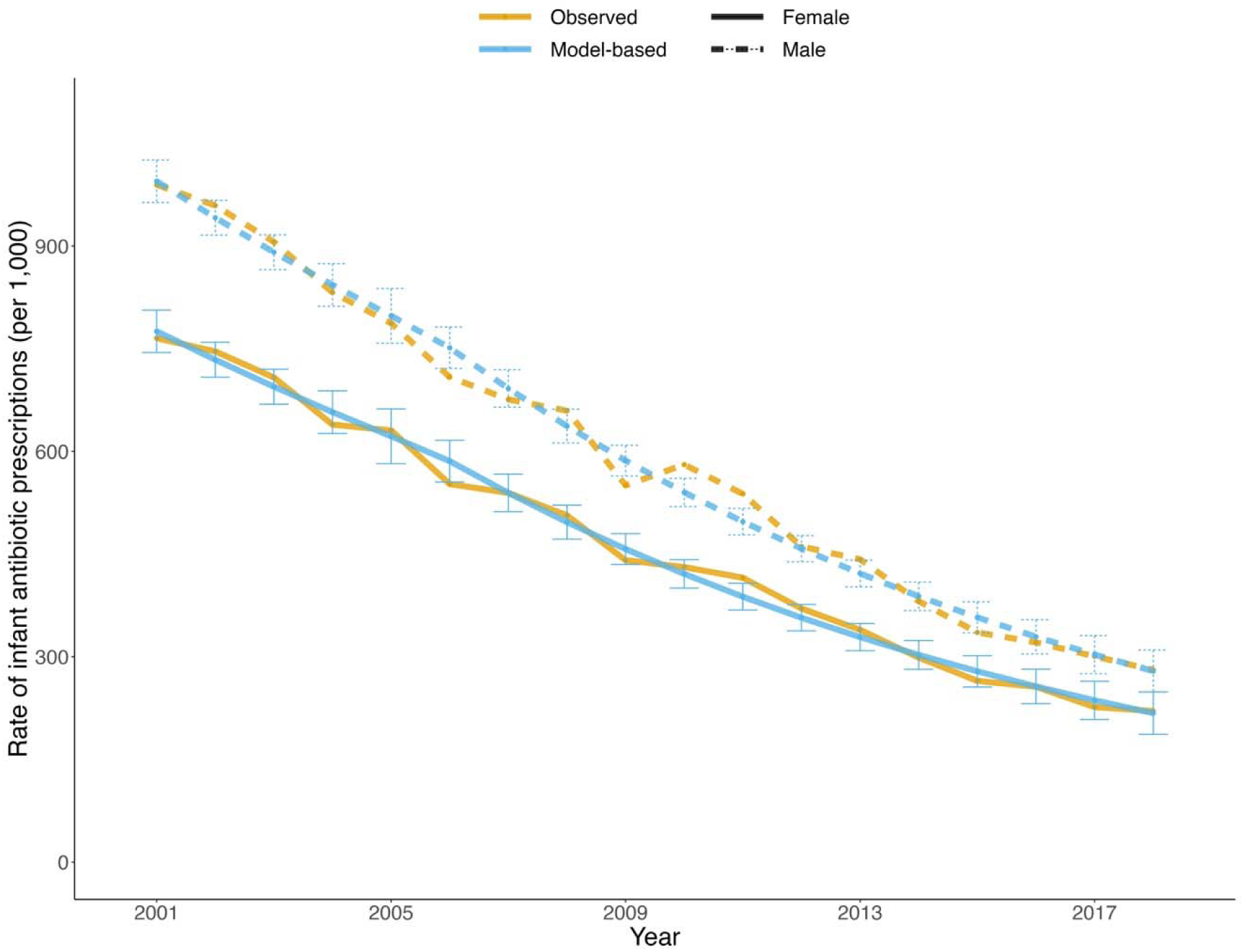
Observed (orange) and model-based (blue) trends in annual rate of infant antibiotic prescriptions by sex (solid: female; dotted: male) from 2001 to 2018 in British Columbia. The error bars indicate the 95% confidence intervals of the model-based values.

### 3.2 Dose-response relationship between infant antibiotic use and childhood asthma development

After examining the systematic review by Duong et al. (8) and the literature review of subsequent studies (**Supplementary Materials Section 1**), we found 6 studies that met our inclusion criteria (**Figure 2** (37) and **Supplementary Materials Section 3**). The studies were from Canada, Japan, Poland, South Korea, and the United States of America (11,38–42). Publication years ranged from 2004 to 2020. The most recently published study reported on a prospective cohort in which asthma diagnosis was confirmed by a physician (11). One study reported on a cross-sectional cohort in which asthma diagnosis was based on self-report (41). The remaining four studies reported on retrospective cohorts in which asthma diagnosis was ascertained by international classification of disease (ICD-9 and ICD-10) codes (38–40,42).

**Figure 2.**
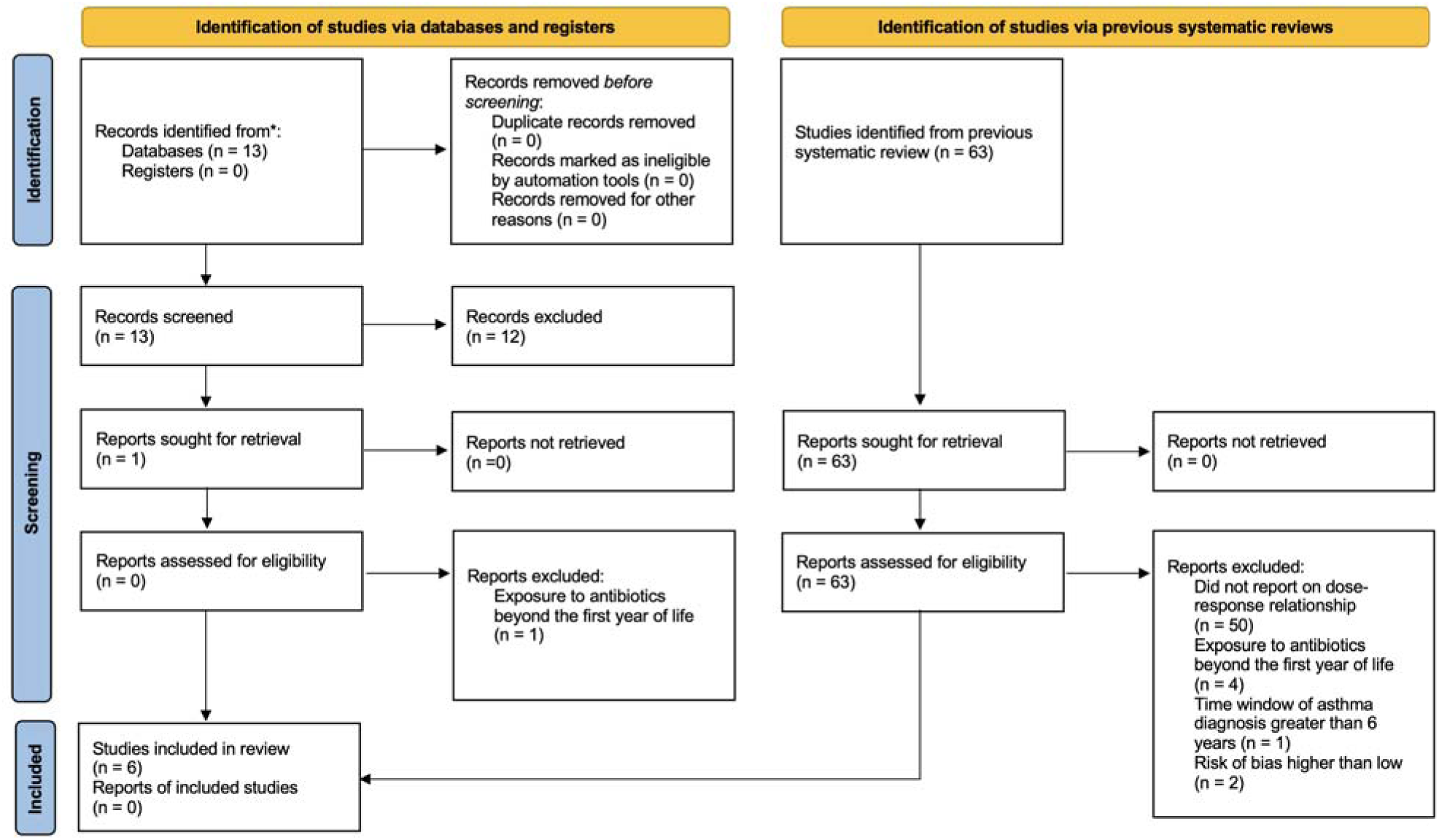
Flow diagram for meta-analysis. *MEDLINE Ovid was used to identify the records (see Supplementary Materials Section 3 for details).

Figure 3 illustrates a dose-response relationship between the number of courses of antibiotics prescribed in the first year of life and asthma prevalence at different ages (3 years was the minimum age, as LEAP does not assign asthma attributes to children below this age) from the Bayesian meta-analysis (parameter estimates are provided in **Table 1**). There was a clear dose-response pattern, with the effect being higher with higher levels of antibiotic exposure (adjusted odds ratio of the dose effect: 1.05; 95% CI: 1.03–1.07) but attenuating over time (adjusted odds ratio of the ageing effect: 0.79; 95% CI: 0.77–0.82). Given the substantial attenuation at 7 years of age, we conservatively assumed the association is completely diminished beyond 7 years of age.

**Figure 3.**
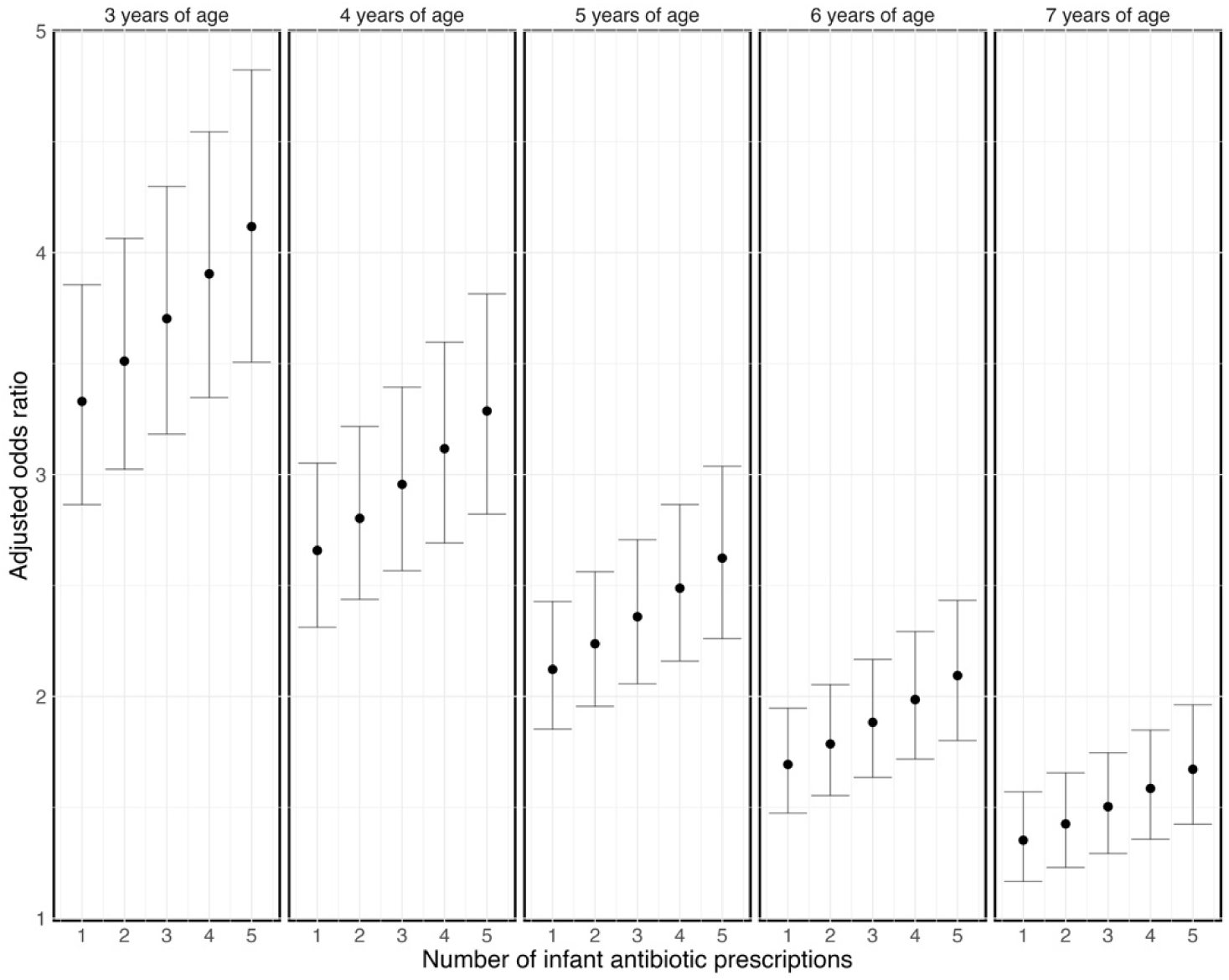
Dose-response relationship between infant antibiotic prescriptions in the first year of life and childhood asthma prevalence at different ages for Canada. The error bars indicate the 95% prediction intervals.

**Table 1.** Parameter estimates from the meta-regression of asthma prevalence. *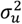 refers to the study-to-study variance component (see **Supplementary Materials Section 3** for details).

| Parameter | Estimate | 95% credible interval |
| --- | --- | --- |
| $\beta_0$ | 1.71 | (0.77, 2.54) |
| $\beta_{Canada}$ | 0.12 | (-0.69, 1.06) |
| $\beta_{age}$ | -0.23 | (-0.26, -0.20) |
| $\beta_{dose}$ | 0.05 | (0.03, 0.07) |
| $\sigma_u^2*$ | 0.93 | (0.48, 1.99) |

### 3.3 Population impact of counterfactual infant antibiotic exposure scenarios

The modeled antibiotic exposure trends by sex are presented for each scenario in Figure 4. The model produced asthma incidence and prevalence rates as expected under the base (factual) scenario (see **Supplementary Materials Section 4**). Furthermore, the decrease of 26.4% (MC SD: 0.2%) in asthma incidence among children of 1–4 years of age under the base scenario between 2001 and 2014 closely matched the reported value (26.0%) in the same age group in a similar period between 2000 and 2014 in BC in another independent study (11).

**Figure 4.**
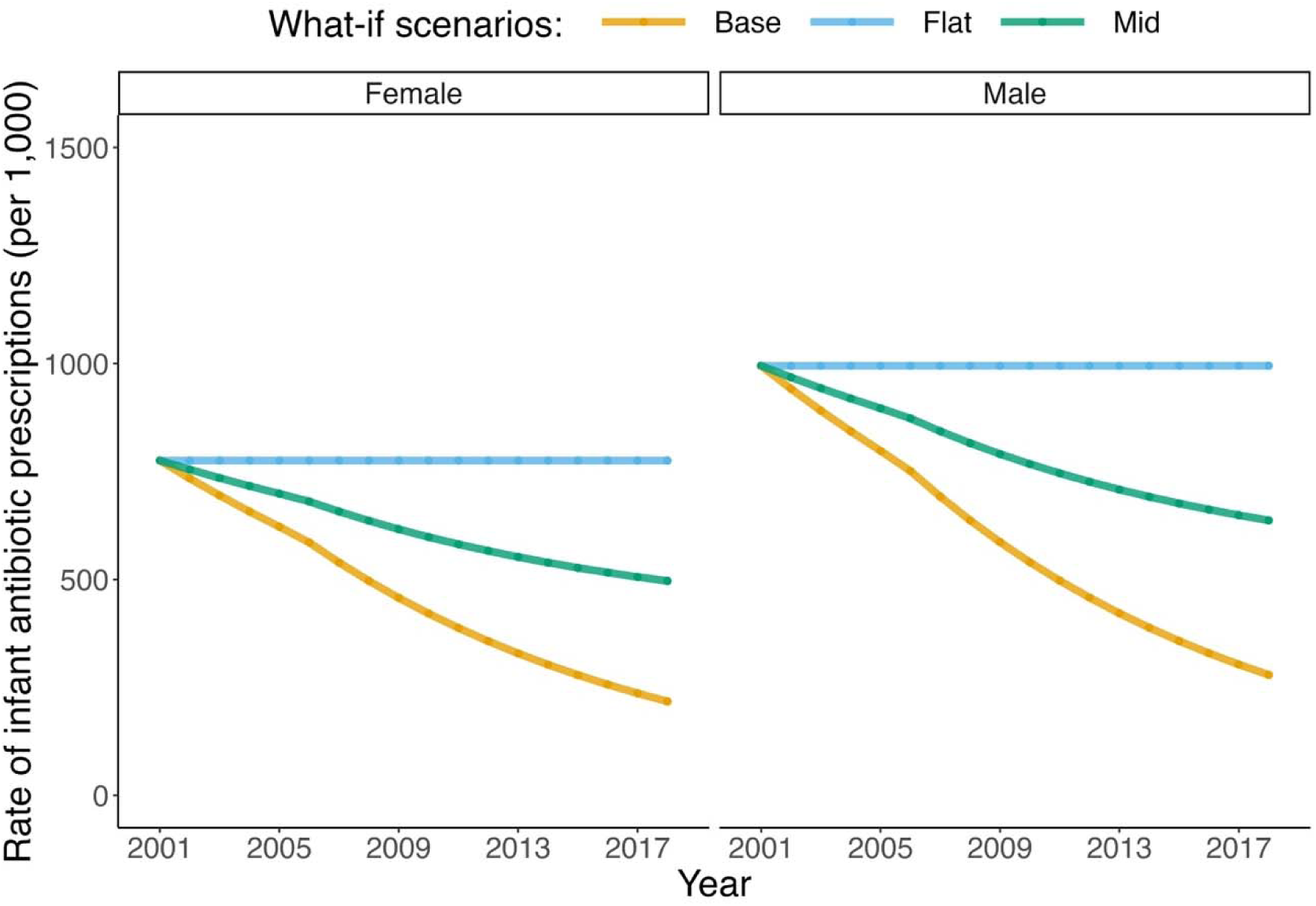
Trends in antibiotic exposure in infants for the three modeled scenarios. *Scenario 1 (base): smoothed trend fitted to observed data in the antibiotic prescription rate in British Columbia *Scenario 2 (flat): trend in the prescription rate if it remained as the smoothed values in 2001 *Scenario 3 (mid): trend in the prescription rate if it declined by half of the smoothed trend.

In Scenario 1 (base scenario), the model simulated an average (across 100 runs) of 1,982,861 (MC SD: 4,161) person-years with asthma, an average of 183,392 (MC SD: 417) cumulative asthma incident cases, and an average of 383,072 (MC SD: 1,129) cumulative exacerbations for the pediatric population (**Table 2**). In comparison, Scenario 2 resulted in estimated 1.9–6.1%, depending on the outcome, relative increases in asthma outcomes during the study period. In absolute terms, if no decline in antibiotic exposure had happened, there would have been an additional 37,213 (MC SD: 5,370) person-years with asthma, 10,053 (MC SD: 558) asthma incident cases, and 23,280 (MC SD:1,455) pediatric exacerbations between 2001 and 2018 (Figure 5). The younger age groups (3–4 and 5–9) were responsible for the majority of the excess burden: 85.4% for cumulative person-years with asthma, 100.0% for cumulative asthma incident cases, and 98.2% for cumulative asthma exacerbations.

**Figure 5.**
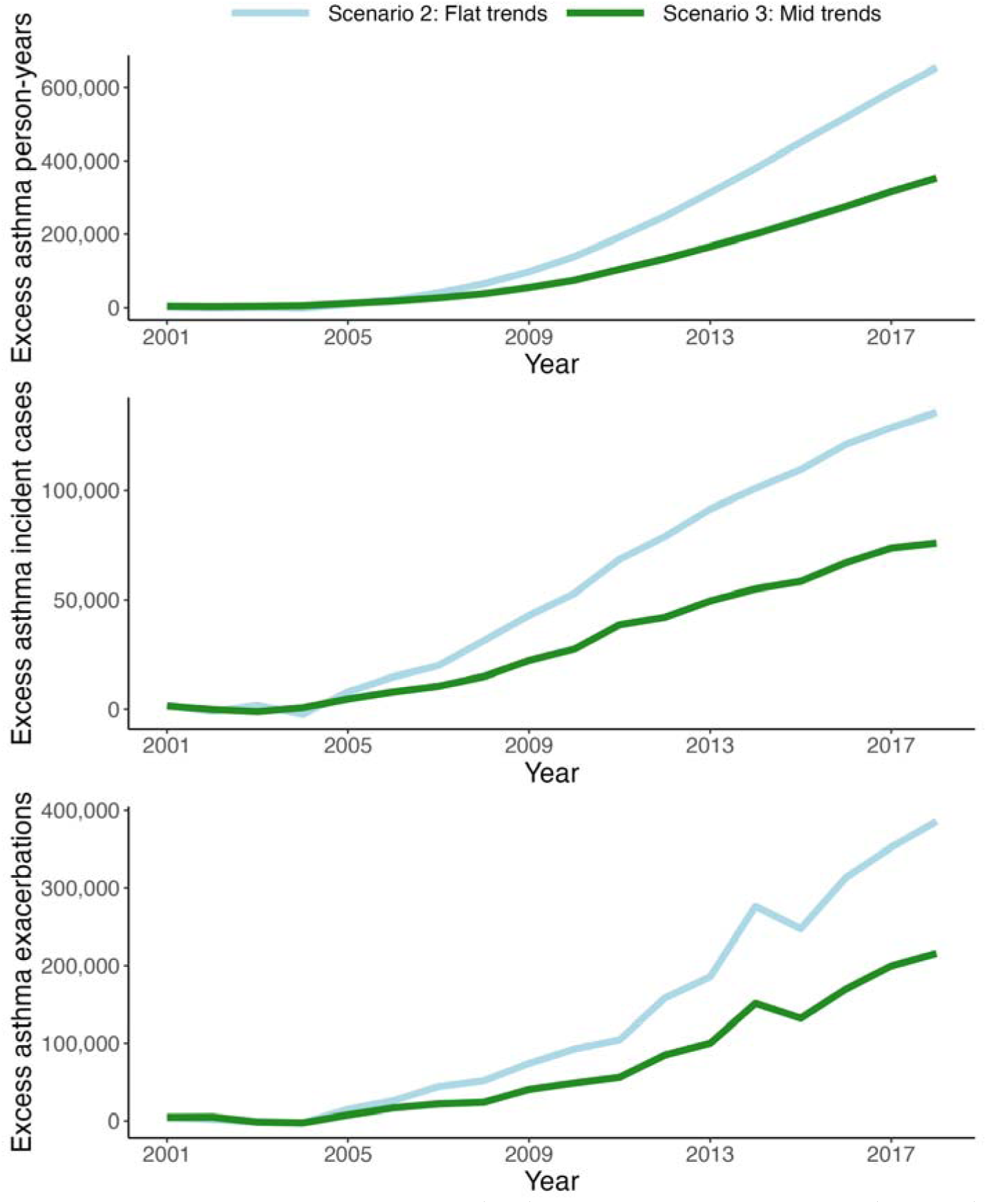
Excess number of asthma person-years (top), asthma incident cases (middle), and exacerbations (bottom) in the counterfactual scenarios (flat and mid) compared with the base scenario for the pediatric population (3–18 years of age). *Scenario 1 (base): smoothed trend fitted to observed data in the antibiotic prescription rate in British Columbia *Scenario 2 (flat): trend in the prescription rate if it remained as the smoothed values in 2001 *Scenario 3 (mid): trend in the prescription rate if it declined by half of the smoothed trend.

**Table 2.**
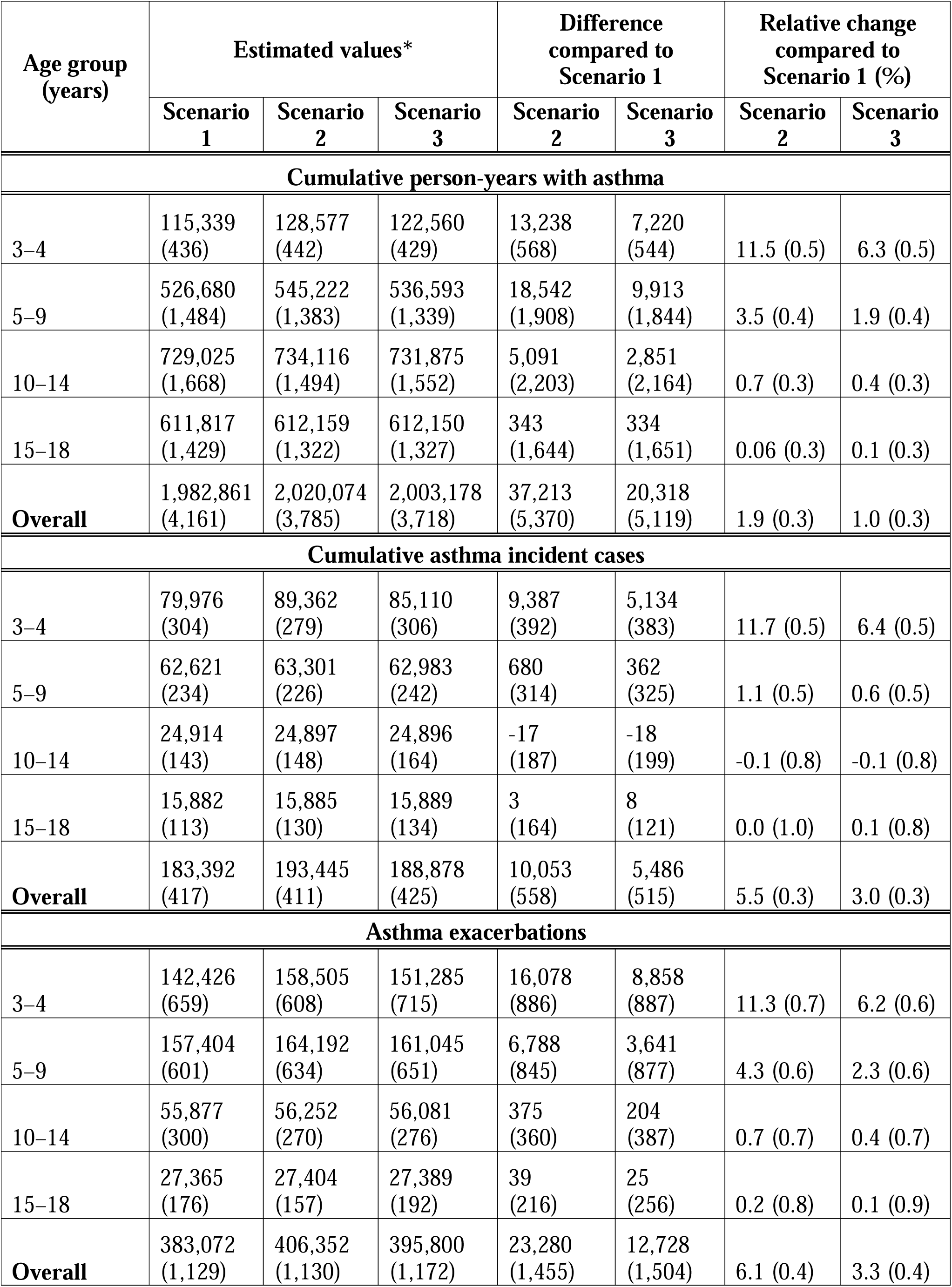
Evaluation of different trends in infant antibiotic use on asthma-related outcomes for the entire pediatric population (3–18 years of age) and by the age groups between 2001 and 2018. The values in the brackets are the Monte Carlo SD (across 100 runs). *Average across 100 runs. **Scenario 1 (base): fitted observed trends in the antibiotic prescription rate in British Columbia **Scenario 2 (flat): trends in the prescription rate that remained as observed in 2001 **Scenario 3 (mid): trends in the prescription rate that declined by half of the fitted observed

| Age group<br>(years) | Estimated values* |  |  | Difference<br>compared to<br>Scenario 1 |  | Relative change<br>compared to<br>Scenario 1 (%) |  |
| --- | --- | --- | --- | --- | --- | --- | --- |
|  | Scenario<br>1 | Scenario<br>2 | Scenario<br>3 | Scenario<br>2 | Scenario<br>3 | Scenario<br>2 | Scenario<br>3 |
| **Cumulative person-years with asthma** |  |  |  |  |  |  |  |
| 3–4 | 115,339<br>(436) | 128,577<br>(442) | 122,560<br>(429) | 13,238<br>(568) | 7,220<br>(544) | 11.5 (0.5) | 6.3 (0.5) |
| 5–9 | 526,680<br>(1,484) | 545,222<br>(1,383) | 536,593<br>(1,339) | 18,542<br>(1,908) | 9,913<br>(1,844) | 3.5 (0.4) | 1.9 (0.4) |
| 10–14 | 729,025<br>(1,668) | 734,116<br>(1,494) | 731,875<br>(1,552) | 5,091<br>(2,203) | 2,851<br>(2,164) | 0.7 (0.3) | 0.4 (0.3) |
| 15–18 | 611,817<br>(1,429) | 612,159<br>(1,322) | 612,150<br>(1,327) | 343<br>(1,644) | 334<br>(1,651) | 0.06 (0.3) | 0.1 (0.3) |
| **Overall** | 1,982,861<br>(4,161) | 2,020,074<br>(3,785) | 2,003,178<br>(3,718) | 37,213<br>(5,370) | 20,318<br>(5,119) | 1.9 (0.3) | 1.0 (0.3) |
| **Cumulative asthma incident cases** |  |  |  |  |  |  |  |
| 3–4 | 79,976<br>(304) | 89,362<br>(279) | 85,110<br>(306) | 9,387<br>(392) | 5,134<br>(383) | 11.7 (0.5) | 6.4 (0.5) |
| 5–9 | 62,621<br>(234) | 63,301<br>(226) | 62,983<br>(242) | 680<br>(314) | 362<br>(325) | 1.1 (0.5) | 0.6 (0.5) |
| 10–14 | 24,914<br>(143) | 24,897<br>(148) | 24,896<br>(164) | -17<br>(187) | -18<br>(199) | -0.1 (0.8) | -0.1 (0.8) |
| 15–18 | 15,882<br>(113) | 15,885<br>(130) | 15,889<br>(134) | 3<br>(164) | 8<br>(121) | 0.0 (1.0) | 0.1 (0.8) |
| **Overall** | 183,392<br>(417) | 193,445<br>(411) | 188,878<br>(425) | 10,053<br>(558) | 5,486<br>(515) | 5.5 (0.3) | 3.0 (0.3) |
| **Asthma exacerbations** |  |  |  |  |  |  |  |
| 3–4 | 142,426<br>(659) | 158,505<br>(608) | 151,285<br>(715) | 16,078<br>(886) | 8,858<br>(887) | 11.3 (0.7) | 6.2 (0.6) |
| 5–9 | 157,404<br>(601) | 164,192<br>(634) | 161,045<br>(651) | 6,788<br>(845) | 3,641<br>(877) | 4.3 (0.6) | 2.3 (0.6) |
| 10–14 | 55,877<br>(300) | 56,252<br>(270) | 56,081<br>(276) | 375<br>(360) | 204<br>(387) | 0.7 (0.7) | 0.4 (0.7) |
| 15–18 | 27,365<br>(176) | 27,404<br>(157) | 27,389<br>(192) | 39<br>(216) | 25<br>(256) | 0.2 (0.8) | 0.1 (0.9) |
| **Overall** | 383,072<br>(1,129) | 406,352<br>(1,130) | 395,800<br>(1,172) | 23,280<br>(1,455) | 12,728<br>(1,504) | 6.1 (0.4) | 3.3 (0.4) |

As expected, the relative increase in asthma outcomes (1.0%–3.3% depending on the outcome) was smaller for Scenario 2. In absolute terms, there were an additional 20,318 (MC SD: 5,119) person-years with asthma, 5,486 (MC SD: 515) asthma incident cases, and 12,728 (MC SD: 1,504) exacerbations during the study period compared to the base scenario (**Table 2** and Figure 5).

Similarly, the younger age groups (3–4 and 5–9) were mainly responsible for the excess burden: 84.3% for cumulative person-years with asthma, 100.0% for cumulative asthma incident cases, and 98.2% for cumulative asthma exacerbations.

## 4 Discussion

In this counterfactual population-level impact analysis, we used real-world trends from population-based data, quantitative evidence synthesis, Bayesian meta-analysis, and simulation modeling to quantify the population-level impact of changes in infant antibiotic exposure on the burden of asthma in a well-defined population over an eighteen-year period (2001–2018). The observed decline in antibiotic use was juxtaposed to two counterfactual scenarios, modeling the trends in antibiotic exposure in the first year of life that were different than the sustained decline observed during this period. We found that our projected results based on observed exposure trends closely matched the decrease in asthma incidence among children of 1–4 years of age between 2000 and 2014 in BC reported from a separate study (11), further validating the use of the policy model in our study. We estimated that if infant antibiotic use had not been reduced from 2001 levels, there would have been excess 37,213 person-years with asthma in the pediatric population in this period. Putting this number in perspective, assuming annual direct medical costs of $550 (in 2024 Canadian dollars) for each person with asthma (43), BC has saved approximately $20 million on the burden of asthma in the pediatric population alone by reducing unnecessary use of antibiotics among infants during the study period (of note, the savings will higher if we include costs of antibiotic prescriptions).

These results can be seen as a prime example of far-reaching implications of reduction in antibiotic exposure in infants. Asthma is one of the manifestations of allergic diseases, whose risk is postulated to be affected by changes in gut microbiota (10). Considering the broader allergic conditions is likely to result in higher estimated benefit from reducing unnecessary antibiotic exposure. As well, the maximum age within which asthma outcomes were modeled in this study was 18 years (for infants born in 2001). For infants born later during the study period this time was shorter. A prevented case of asthma averts asthma-related burden throughout the life of the individual, translating to avoided exacerbations, reduction in quality of life due to symptoms, missed school days among children, and work productivity loss among adults for many years. As such, following the infant cohorts born within the study period into the future is likely to substantially increase the estimated benefits of antibiotic exposure reduction.

Already, without accounting for such benefits, interventions and policies aimed at reducing antibiotic exposure are found to be effective and most likely cost-saving (2,44). A systematic review of 52 studies on the evaluation of antimicrobial stewardship programs revealed that they were associated with a 10% reduction in antibiotic prescriptions. The association was stronger for the low- and middle-income countries (an average decrease of 30%) and for pediatric patients (an average decrease of 21%), implying antimicrobial stewardship programs can be an effective intervention to deal with overuse of antibiotics. However, effectiveness alone, without considering costs, cannot guide decision-making. The Organization for Economic Co-operation and Development recently evaluated antimicrobial resistance control policies, including antimicrobial stewardship programs, across all its member countries (44). They used a microsimulation model that incorporated the impact of antibiotics on antimicrobial-resistant infections and modeled counterfactual scenarios to compare with a “no intervention” base scenario. They found that the stewardship programs would be cost-saving with a probability of at least 70%, cost-effective (at a threshold of US$50,000 per quality-adjusted life year) with a probability of at least 20%, and there was a <10% change of being not cost-effective. The chance of being cost-effective and cost-saving was likely underestimated, as the above-mentioned analysis did not consider the impact of antibiotic exposure on the burden of other diseases, such as asthma.

Ours was a simulation study on the benefit of reducing unnecessary antibiotic use on asthma-related outcomes based on the latest available evidence linking the two. In Canada, BC has the lowest antibiotic prescription rate among children. While this does not imply that BC has the lowest rate of unnecessary antibiotic prescriptions, the decrease in antibiotic use is attributed in part to an antimicrobial stewardship program in BC (24,25). As such, this potentially indicates more room for improvement in other provinces and territories (45). This will help Canada break stagnant trends in asthma prevalence and severe asthma exacerbations requiring inpatient care (46,47). Higher benefits are expected for many other jurisdictions, including low- and middle-income countries, many of which are observing an upward trend in antibiotic use among children under 5 years of age (48). Urgent initiatives involving collaboration among healthcare professionals, researchers, and policymakers are required to change the course of this trend and cope with a high proportion of overuse of antibiotics in both low- and middle-income countries and high-income countries (49,50).

This study was not an analysis of any specific intervention for the reduction of antibiotic exposure. Instead, we focused on quantifying the current evidence on the association between antibiotic exposure and the population-level burden of asthma. Evaluating the outcomes of a specific intervention (e.g., implementing an antibiotic stewardship program in a jurisdiction) requires evidence on its resource implication, the effectiveness of the intervention on antibiotic use, as well as the multifaceted downstream implication of such change in the exposure. Dimensions of benefit from reducing unnecessary antibiotic use should be first studied in detail, as was done here for asthma, based on systematic and quantitative evidence synthesis and outcome modeling. Studies like ours, conducted for other conditions that might be affected by changes in antibiotic exposure, will provide evidence for a future meta-modeling study, collating all facets of the benefits and harms of exposure reduction initiatives. This will provide a complete picture of the effects of a specific intervention.

Our study has several strengths. Our estimation of infant antibiotic exposure was obtained from comprehensive, population-based data from the entire target population of a well-defined geographic area and health jurisdiction, without uncertainty due to sampling or bias due to representativeness. We used Bayesian meta-analysis to quantify high quality evidence from multiple different studies on the nuances of the dose-response association of primary interest. This is in contrast with a data-driven counterfactual analysis (which would have used the same data for both trend analysis of antibiotic exposure and estimation of dose-response association). The latter would restrict the evidence to one dataset. Further, given the nature of the data (administrative), controlling for potential confounding variables would have been difficult. Instead, we reconciled evidence on the trends observed from the population-based data with quantitative meta-analysis of all available evidence, and used computer modeling techniques to propagate such evidence to change in asthma-related outcomes. Thus, simulation modeling allows for addressing the limitations of a single, static population-based study through evidence synthesis and various ‘what-if’ scenarios (51). Of note, the policy model for asthma used in this work has been developed independently of this study (as a multi-purpose platform for policy analysis of asthma-related interventions in Canada) and has undergone rigorous analysis and independent validation studies (34).

The limitations of this study should also be acknowledged. A critical piece of evidence underlying the asthma model is the dose-response relationship between infant antibiotic use and childhood asthma prevalence that we estimated through the meta-analysis. We found a negative association that decayed over time (Figure 3). While the overall risk of bias was low in the included studies, the extent to which this association can be interpreted causally depends on the degree to which the included studies successfully controlled for confounding effects. In particular, potential biases due to confounding by indication or reverse causation were directly addressed in two of the included studies by excluding children who received antibiotics for respiratory symptoms or were diagnosed with asthma in the first year of life (11,40). The remaining four studies could not fully account for confounding by indication or reverse causation (38,39,41,42). We note that in our case, the original dose-response association of the Canadian study (11) was slightly attenuated after pooling evidence from other studies through the meta-analysis. Moreover, due to a lack of evidence in the literature, examining whether the dose-response relationship holds for antibiotic exposure beyond the first year of life is difficult. Evidence from mechanistic studies suggests that the critical window for gut microbiome maturation appears to be within the first 100 days of life (52,53). Not modeling any effect beyond one year therefore seems justified. This conservative assumption means any effect beyond the first year of life will increase the benefits observed in this study. However, in general, more robust evidence is needed to better understand the causal relationship on early life antibiotic exposure and the risk of asthma.

Another limitation is the difficulty in confirming asthma diagnosis in children under 6 years of age (54). Our decision to model asthma cases in that age group (specifically 3 years or older) was to reflect and incorporate current labeling of health utilization related to asthma by healthcare systems. In Canada, asthma cases in the administrative health databases are identified based on a validated case definition (55). Of note, misdiagnosis and remission are accounted for in the LEAP model in order to match asthma prevalence, which rises in early childhood and declines during adolescence (see **Supplementary Figure 3**). Regarding the meta-analysis, three included studies examined the asthma diagnosis in children under 6 years of age (11,38,42). Yoshida et al. used a conservative approach based on a typical diagnostic code case definition combined with confirmed use of ICS and controller medications, likely underestimating asthma cases (38). In contrast, in the studies by Patrick et al. (11) and Celedon et al. (42), asthma diagnosis was confirmed by physicians (which is the recommended approach by the Canadian Thoracic Society (56)). These studies acknowledged the difficulty and potential misclassification of asthma diagnosis in young children despite their efforts. Last but not least, the LEAP model in its current implementation does not consider uncertainty in the evidence. Further, we did not evaluate costs for this study. It is obvious from our results that costs associated with antibiotic use and asthma will both decrease with reduced unnecessary antibiotic exposure (whereas costs will increase if necessary antibiotics are wrongly reduced).

There are several areas for future research. Modeling studies are required for other conditions that might be affected by antibiotic exposure. Specific programs and interventions aimed at reducing unnecessary antibiotic exposure should be evaluated in dedicated analyses, which should also incorporate costs to guide policymaking. Further, there is now robust evidence indicating that the effect of antibiotic use on allergic disease is mediated via the gut microbiota (10,11). We note that another modifier of gut microbiota is breastmilk exposure (57,58). Recent evidence suggests that breastmilk works as an effect-modifier between prenatal and infant antibiotic use and childhood asthma development (17,59). Therefore, a joint intervention targeting infant antibiotic overuse and promoting breastfeeding (particularly where antibiotic exposure is necessary in infancy) can lead to more effective prevention against childhood asthma development. The outcome of such policies should be investigated in the future.

In summary, this study quantified the far-reaching benefits of reducing unnecessary antibiotic use on the burden of asthma. Our findings present compelling evidence to urge jurisdictions with overuse of antibiotics in infants and high prevalence of childhood asthma to implement antimicrobial stewardship initiatives. Incorporating the life-long impact on the burden of asthma and potentially other diseases will increase the value of interventions addressing the overuse of antibiotics in infancy. Future work should incorporate utility and costs to provide a measure on the value-for-money potential of such interventions.

## Author Contributions

TYL: Data curation, Formal analysis, Methodology, Software, Visualization, Writing – original draft. JP: Conceptualization, Methodology, Supervision, Writing – review & editing. AS: Data curation, Writing – review & editing. FM: Data curation, Conceptualization, Writing – review & editing. SET: Funding acquisition, Writing – review & editing. HL: Conceptualization, Writing – review & editing. DMP: Data curation, Conceptualization, Writing – review & editing. JJC: Writing – review & editing. KMJ: Writing – review & editing. MS: Conceptualization, Methodology, Funding acquisition, Supervision, Writing – review & editing.

## Acknowledgments

We would like to acknowledge the Community Antimicrobial Stewardship research team at BC CDC (http://www.bccdc.ca/our-services/programs/community-antimicrobial-stewardship) for their feedback and assistance with access to the data on antibiotic prescriptions.

Access to data provided by the Data Stewards is subject to approval but can be requested for research projects through the Data Stewards or their designated service providers. The following data sets were used in this study: Chronic Disease Registry and Pharmanet. All inferences, opinions, and conclusions drawn in this publication are those of the author(s), and do not reflect the opinions or policies of the Data Steward(s).

## Data Availability Statement

The datasets analyzed and generated for this study can be found in the GitHub repository: https://github.com/tyhlee/ImpactInfantAbxAsthma.

