## Supplementary Material for "Impact analysis of infant antibiotic exposure on the burden of asthma: a simulation modeling study"

1. **Literature review**

We performed a literature review to identify any relevant studies published since 2021. On September 29, 2023, MEDLINE was searched using the Ovid interface. The search terms and the resulting number of publications are provided in Supplementary Materials Table 1. A total of 13 studies were identified with the search terms, and one study(1) was found to be relevant based on titles and abstracts. However, that study did not meet our inclusion criteria (the exposure timing was the first two years of life)

|  | **Search terms** | **Number of publications** |
| --- | --- | --- |
| 1 | exp Asthma/ | 143,117 |
| 2 | (antibiotic or antibiotics or (antibacterial* or anti-bacterial* or bacteriocid* or antimicrobial* or anti-microbial* or ntimicrobial* or anti-mycrobial* or antiinfective or anti-infective)).mp. [mp=title, book title, abstract, original title, name of substance word, subject heading word, floating sub-heading word, keyword heading word, organism supplementary concept word, protocol supplementary concept word, rare disease supplementary concept word, unique identifier, synonyms, population supplementary concept word, anatomy supplementary concept word] | 867,553 |
| 3 | exp *Anti-Bacterial Agents/ or exp Anti-Bacterial Agents/im, tu or *anti-infective agents/ | 659,218 |
| 4 | 2 or 3 | 1,107,314 |
| 5 | 1 and 4 | 2,598 |
| 6 | limit 5 to yr=”2021 – 2024” | 228 |
| 7 | limit 6 to (structured abstracts and full text) | 13 |

**Supplementary Table 1.** Search terms and results using the MEDLINE Ovid.

2. **Midpoint approximation**

We investigated whether our single-time midpoint approximation to a short age range of asthma diagnosis was plausible. We considered the binary exposure case (no antibiotics versus at least one course of antibiotics) in the first year with a simple data generating mechanism. The probability of having asthma for a given age and exposure is provided by the following equation:

Logit(asthma prevalence | age, exposure) = β_0_ + β_age_*age + β_exposure_*exposure+β_age:exposure_*age:exposure

We set the parameter values such that the probability decayed over time and the effect of antibiotic exposure in the first year diminished completely at age 10 (see **Supplementary Figure 1**). With the sample size of n=1,000,000 individuals, we considered three ranges of 3 (e.g., 5–7 years of age), 5 (e.g., 1–5 years of age), and 7 (4–10 years of age) in years. For each individual, we randomly assigned age between 1 and 10 years with a uniform distribution, exposure status with a Bernoulli distribution with probability of 0.40, and asthma status with β_0_=-3.5, β_age_=-0.01, β_exposure_=0.8, and β_age:exposure_=-0.08.

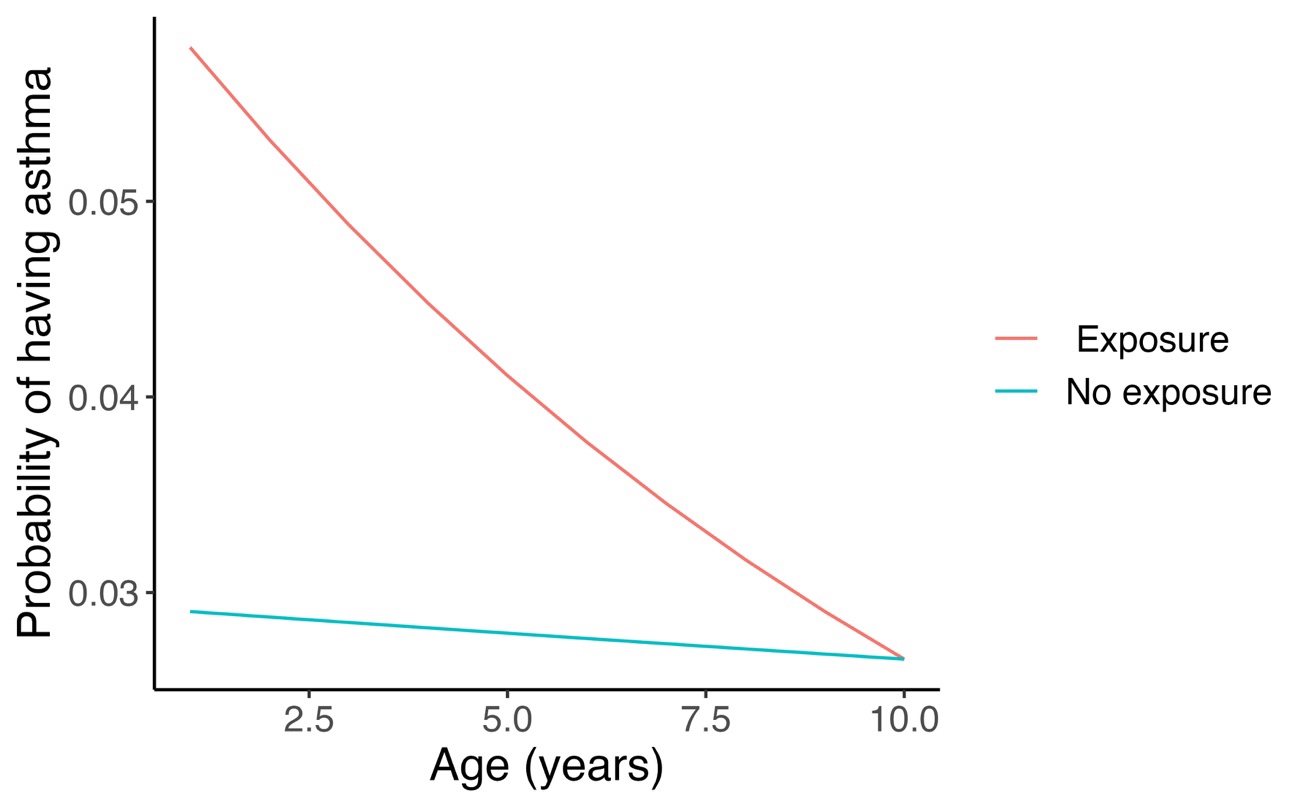

**Supplementary Figure 1.** Probability of having asthma for the simulation study of the midpoint approximation.

For each age rage, we computed the age-specific OR in the midpoint year and the overall OR using the simulation results (**Supplementary Table 2**). The midpoint approximation seems to be close to the overall OR (over all the ages in the time range) for these short ranges.

| **Age range (years)** | **Length of the range (years)** | **Midpoint OR** | **Overall OR** |
| --- | --- | --- | --- |
| 1 – 3 | 3 | 1.91 | 1.91 |
| 2 – 4 | 3 | 1.73 | 1.74 |
| 3 – 5 | 3 | 1.58 | 1.64 |
| 4 – 6 | 3 | 1.61 | 1.51 |
| 5 – 7 | 3 | 1.34 | 1.42 |
| 6 – 8 | 3 | 1.31 | 1.28 |
| 7 – 9 | 3 | 1.18 | 1.20 |
| 8 – 10 | 3 | 1.10 | 1.08 |
| 1 – 5 | 5 | 1.73 | 1.78 |
| 2 – 6 | 5 | 1.58 | 1.64 |
| 3 – 7 | 5 | 1.61 | 1.52 |
| 4 – 8 | 5 | 1.34 | 1.40 |
| 5 – 9 | 5 | 1.31 | 1.31 |
| 6 – 10 | 5 | 1.18 | 1.18 |
| 1 – 7 | 7 | 1.58 | 1.65 |
| 2 – 8 | 7 | 1.61 | 1.53 |
| 3 – 9 | 7 | 1.34 | 1.41 |
| 4 – 10 | 7 | 1.31 | 1.30 |

**Supplementary Table 3**. Simulation results on the midpoint approximation.
*OR: odds ratio

The code for the simulation study is included in the appendix of the meta-analysis in the GitHub repository (<https://github.com/tyhlee/ImpactInfantAbxAsthma>).

3. **Details of the meta-analysis**

**Characteristics of the included studies in the meta-analysis**

We provide the summary of characteristics of the six included studies in the meta-analysis.

| **Author** | **Country** | **Study year** | **Study type** | **Asthma diagnosis** | **Age of asthma diagnosis** | **Sample size** | **Number of asthma cases** |
| --- | --- | --- | --- | --- | --- | --- | --- |
| Patrick et al. | Canada | 2020 | Prospective cohort | Care provider | 5 | 1947 | 118 |
| Yoshida et al. | Japan | 2018 | Retrospective cohort | ICD | 2, 5 | 83470 | 3633 |
| Raciborski et al. | Poland | 2012 | Retrospective cohort | Self-reported | 7 | 1330 | 63 |
| Kozyrskyj et al. | Canada | 2007 | Retrospective cohort | ICD | 7 | 13116 | 787 |
| Ahn et al. | Korea | 2005 | Cross-sectional cohort | Self-reported | 9.5 | 25787 | 2354 |
| Celedon et al. | USA | 2004 | Retrospective cohort | ICD | 1.5, 3.5 | 4408 | 323 |

**Supplementary Table 4.** Study characteristics of the included studies in the meta- analysis.

USA: United States of America; ICD: International Classification of Disease codes

**Random-effects meta-regression model**

We fitted a random-effects meta-regression model with age of asthma diagnosis (age) and the number of infant antibiotic prescriptions (dose) as covariates and with adjusted odds ratio (aOR) as the response variable. For study $i,$

$$\log\left( aOR_{i}\left( age,dose \right) \right)\sim Normal\left( \beta_{0}+\beta_{1}age+\beta_{2}dose+u_{i},\sigma_{i}^{2} \right)$$

$$\beta_{0},\beta_{1},\beta_{2} \sim Normal\left( 0,\Sigma\right)$$

$$\Sigma=V\left( L*L^{T} \right)V \left( Cholesky factorization \right)$$

$$u_{i}\sim Normal(0,\sigma_{u}^{2})$$

We used the standard error of $\log\left( aOR_{i}\left( age,dose \right) \right)$ reported in study $i$ as if it were true value of $\sigma_{i}^{2}$. We deemed this approach reasonable, considering the large size of each of the included studies. The priors were specified as follows.

$$\sigma_{u}\sim LogNormal\left( {0,10}^{2} \right)$$

$$V_{ii}\sim Cauchy(0,5)$$

$$L\sim LKJ\_corr\_cholesky(1.0)$$

Note that $LKJ\_corr\_cholesky(1.0)$ implies that the probability density is uniform over all possible correlation matrices (2).

Fitting this model in R Stan provides the posterior distributions of the coefficients along with their summary statistics including their posterior means and credible intervals. Moreover, it provides the posterior prediction of the random-effects term $u_{i}$ for each study $i$ (this was how we obtained $\beta_{Canada}$in Eq. 1). Despite the small sample size of the included studies (n=6), the model was fitted well. For each parameter in **Table 1**, the effective sample size of the Monte Carlo Markov Chain (MCMC) samples was at least 50% (implying at least 5,000 out of 10,000 MCMC samples were independent draws) and the R-hat statistic was equal to 1 (implying the model successfully converged for these parameters).

The R Stan code is provided in the GitHub repository (<https://github.com/tyhlee/ImpactInfantAbxAsthma>).

4. **Validation of the asthma policy model under the base (factual) scenario**

We show three figures for the validation of the asthma policy model under the base (factual) scenario: demographics, asthma incidence and asthma prevalence. For further details, see Lee et al.(3)

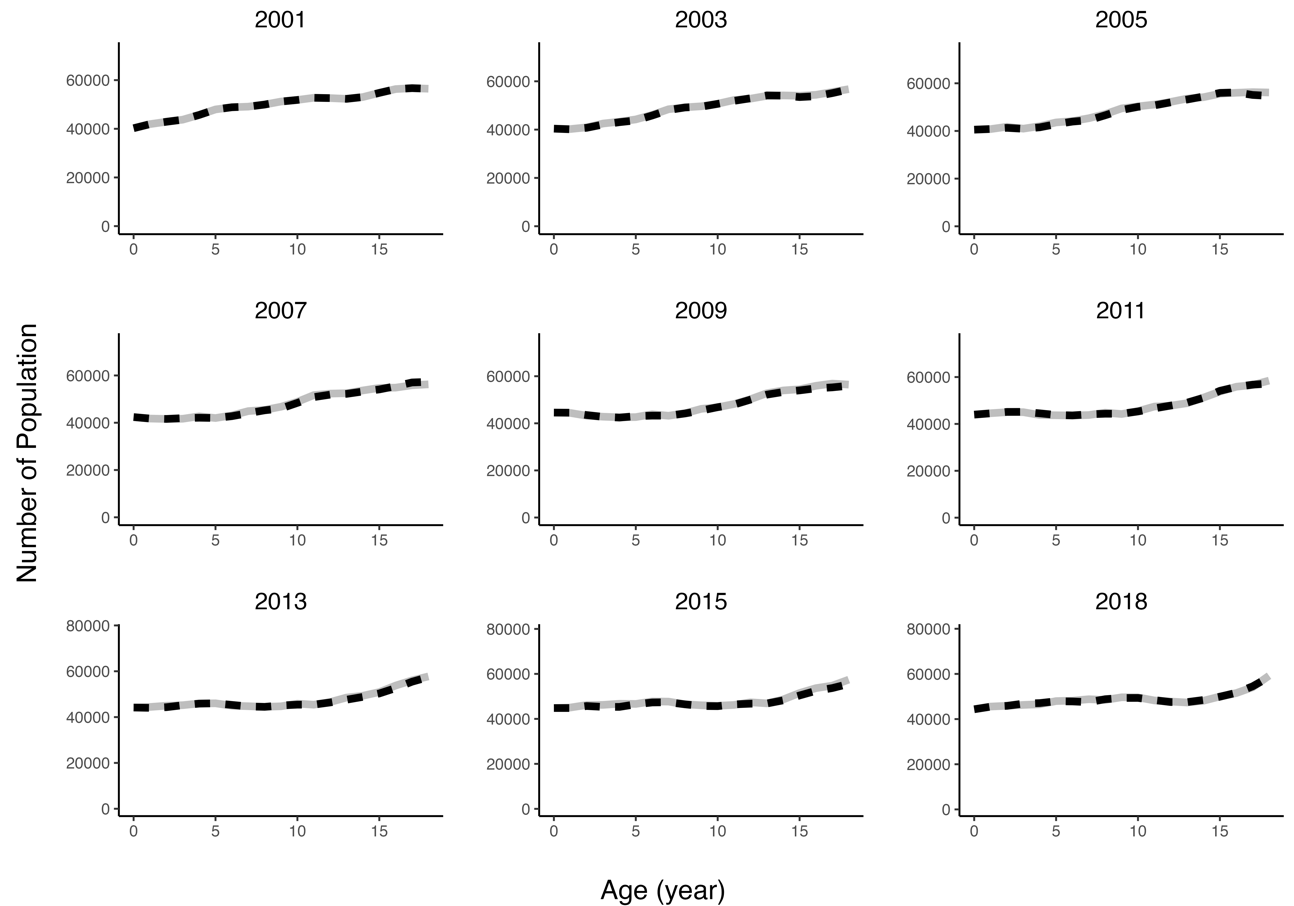
**Supplementary Figure 2**: Population by age across selected years from the model (grey solid) and from Statistics Canada (black dashed).

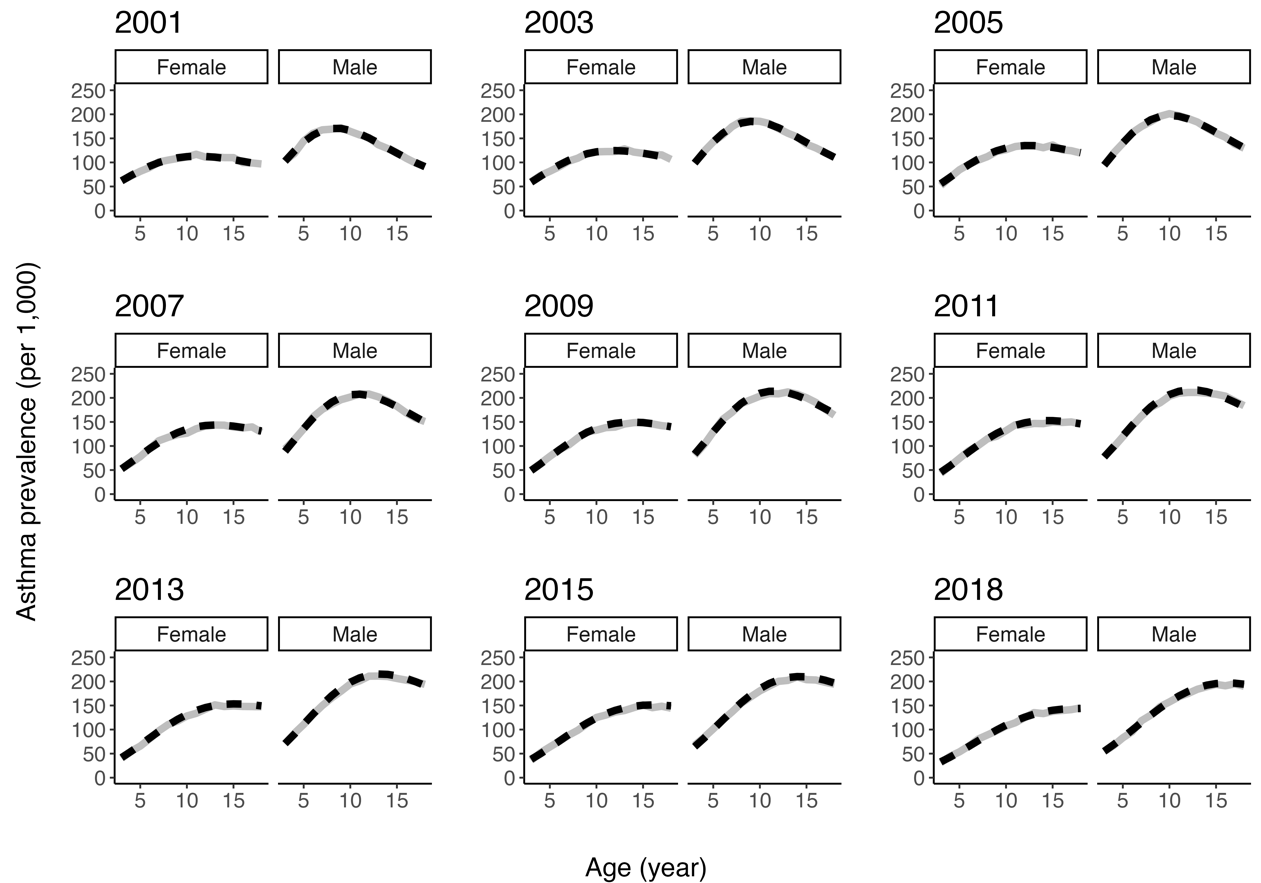
**Supplementary Figure 3**: Asthma prevalence rates per 1,000 general population from the model (solid grey) and estimated (black dashed) under the base scenario.

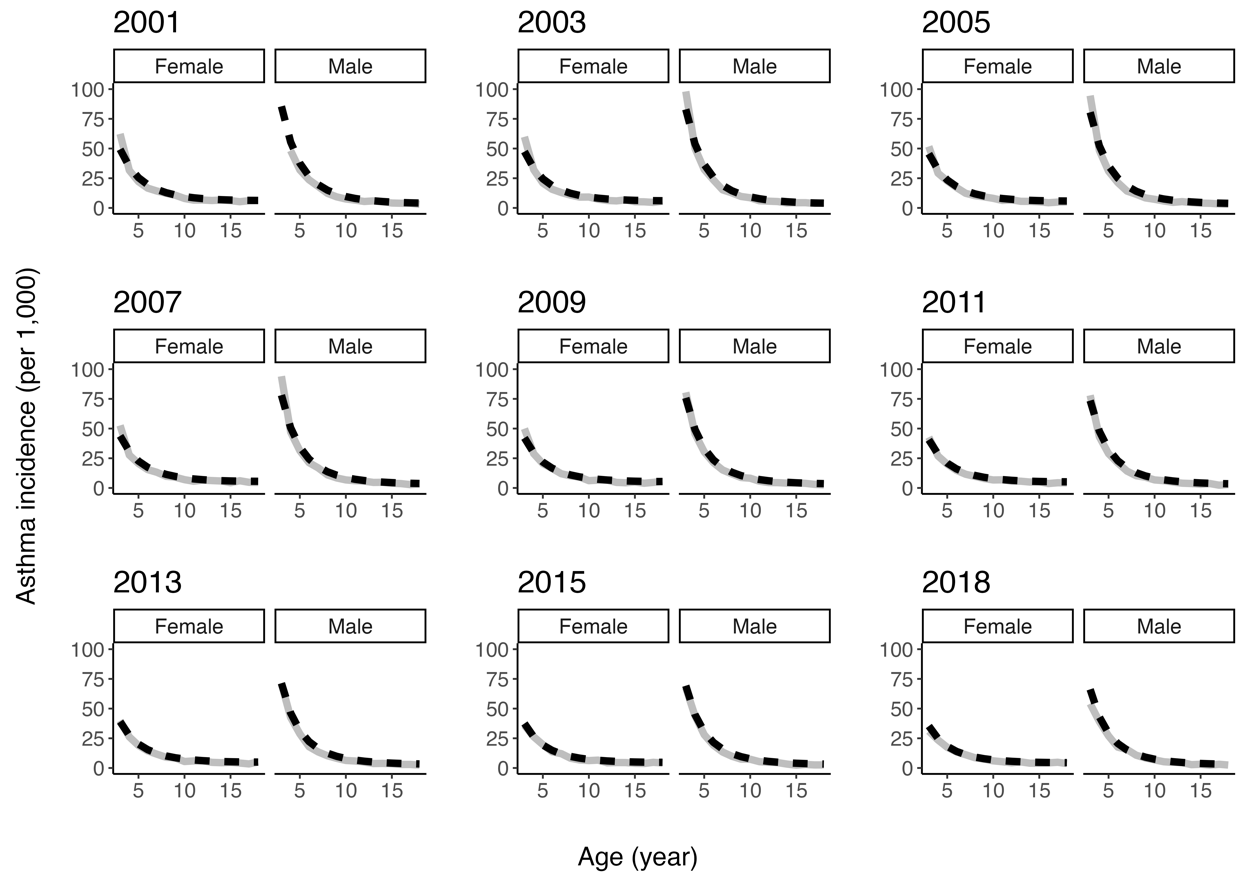

**Supplementary Figure 4**: Asthma incidence rates per 1,000 general population from the model (grey solid) and estimated (black dashed) under the base scenario.
